# Automated vision screening of children using a mobile graphic device

**DOI:** 10.1101/2021.04.29.21255800

**Authors:** Steven A. Kane, Mark Gaspich, Julia Kane, Sarah Weitzman, Albert Hofeldt

## Abstract

**Background/Objective:** Can measures of interocular brightness disparity, acuity, and colour vision identify children with amblyopia?

**Subjects/Methods:** 208 subjects from 3 to 14 years were recruited for a prospective, observational protocol to measure interocular brightness disparity, acuities with and without a pinhole, and colour vision using an iPad. Interocular brightness disparity was assessed as the subject looked through a system of polarizing filters and chose the brighter of two spaceships. The differential brightness of image pairs was varied according to a staircase algorithm until equal brightness was perceived. Acuities were tested with tumbling Es. Colour vision was tested with AO-HRR colour plates. 2 subjects (1%) were later confirmed to have unilateral amblyopia.

**Results:** Binocular brightness balance on the iPad detected both amblyopes and excluded all 202 non-amblyopes, in this study with sensitivity and specificity of 100%. By using 20/30 as cutoff for normal acuity, 1 of the 2 amblyopes was detected and all non-amblyopes were excluded by visual acuity testing with pinhole. The mean difference between iPad and E-Chart visual acuities with pinhole was 0.02 logMAR with limits of agreement from -0.08 to +0.11 logMAR. Colour vision testing with iPad and printed plates gave identical results. Testing times were brief and exit pleasure poll responses were positive.

**Conclusions:** Interocular brightness disparity, acuity, and colour vision can be measured in children as young as 3 years playing a fun game on a mobile graphic device. Interocular brightness disparity may be a sensitive and specific method to detect unilateral amblyopia.

## INTRODUCTION

Amblyopia, an often silent and elusive disease, remains the leading cause of permanent vision loss in children^1^ despite more than a century of interest in vision screening.^2^ Are the screening techniques at fault, are follow up and therapy at fault, or are not enough children being screened for amblyopia? Ideal vision screening would have low rates of false positive and false negative results, low expense, and ready availability. A method having these qualities that could also be administered via telemedicine could improve vision screening in schools and pediatric offices and reach more children who are not being screened for amblyopia.

Relative brightness sense was found to agree closely with the degree of visual acuity impairment in adult subjects with a range of ophthalmic diseases, including amblyopia.^3^ This study investigates the utility of vision screening with a mobile graphic device (iPad) to measure interocular brightness disparity, visual acuity, and colour vision for detecting amblyopia in pediatric subjects in a school setting.

## SUBJECTS AND METHODS

This study utilized a prospective, observational protocol that followed all the tenets of the Declaration of Helsinki and was approved by the Columbia University Institutional Review Board (protocol AAAC0020). 208 children, 121 girls and 87 boys with ages from 3 to 14 years and mean of 7.8 years, were recruited as subjects and tested at school. The protocol measures brightness disparity, acuity, and colour vision with self-tested algorithms running on an iPad with results stored on the device. The age distribution of these subjects was 15.4% (3-5 years of age), 46.6% (6-9 years of age) and 37.9% (10-14 years of age). Their ocular histories were not known beyond the use of spectacles until testing was completed.

To measure brightness disparity, binocular image separation is created by wearing polarizing glasses combined with complementary linear polarizing filters positioned over two vertically aligned spaceships on an iPad screen (Figure 1). Through this polarizing filter arrangement, the right eye views only the bottom spaceship, rivalrous with the black background viewed by the left eye, and the left eye views only the top spaceship, rivalrous with the black background viewed by the right eye. Tops and bottom spaceships are presented with brightness differences ranging from 0.3 to 1.8 log units in increments of 0.3. In response to recorded instruction, the subject identifies and taps the brighter spaceship. The brightness difference of the spaceships and response times are recorded on the device. In response to the subject’s selection of the brighter spaceship, brightness differences of subsequent spaceship pairs are then sequenced within a stepwise, self-tested algorithm until the right-left brightness equality endpoint is crossed and re-crossed. For a normal score the students must achieve a net zero brightness imbalance in 2 of 3 games.

**Figure 1.**
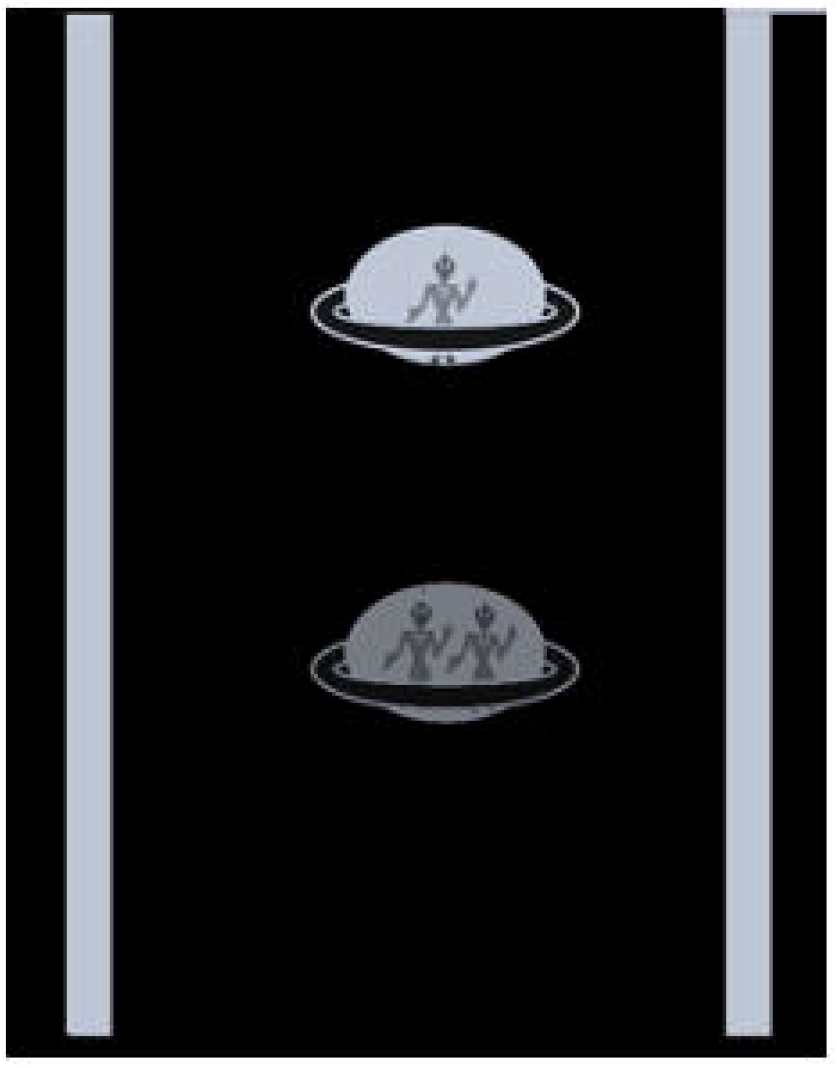
Screenshot of two spaceships presented on an iPad.

iPad visual acuity is based on matching tumbling Es calibrated from 20/400 to 20/20 for a testing distance of 40 cm. A tape measure attached to an iPad stand confirms the testing distance. Reversible spectacles that occlude one eye are worn. Testing begins by the examiner selecting a starting E size, typically 20/60. Two equally sized Es are presented. The student taps YES on the touchscreen when the orientations of the Es are identical and NO when the Es are mismatched. Three correct responses advance the protocol to the next lower line. An incorrect response provides a second chance during presentation of three E pairs of that letter size. Another incorrect response at that E size terminates the self-test. Testing then restarts by the examiner selecting a larger starting E size. The smallest E size with 3 correct responses is recorded as the visual acuity for each eye. When visual acuity measures 20/30 or worse, testing is repeated through a plastic panoramic pinhole (PH) disc containing seven 6 mm opaque rings, each with a 1.0 mm central piercing and each ring margin separated by 1 mm of clear plastic. If the subject fails to match the 20/400 E, the acuity is recorded as less than 20/400. For comparison to distance acuity, the acuities of 63 subjects were also tested with tumbling Es (E-Chart) on a traditional eye chart at 20 feet.

Digital copies of the demonstration and test AO-HRR colour plates are presented on an iPad. Reversible spectacles that occlude one eye are worn. The subject is asked to touch a coloured shape or signify no coloured shape by touching a no colour circle below. If the demonstration plates are correctly identified, the subject qualifies to proceed. The test colour images are then similarly presented in a pseudo-random order and then repeated for the other eye.

Subjects with abnormal results were referred for complete ophthalmological examination if they were not already under care.

## RESULTS

Of the 208 subjects were recruited for testing, 204 subjects were able to complete the protocol to measure interocular brightness disparity, acuities, and colour vision. Except for one amblyope, the visual acuities of the remaining 203 subjects were 20/30 or better in each eye either unassisted, with corrective spectacles, or with the aid of the PH. Four subjects were excluded from the protocol due to either not understanding the visual acuity tests (2 students) or omission of PH acuity testing (2 subjects). The only recruited subject who was unable to successfully play the brightness disparity game was a young child who was also unable to perform the acuity and colour vision tests.

For statistical analysis, acuities were converted to logMAR notation. For the 63 students (126 eyes) tested with both E-Chart and the iPad acuity without the PH, the mean logMAR acuities for E-Chart was 0.11 (standard deviation SD = 0.16) and for iPad acuity was 0.07 (SD = 0.12). With the PH, the mean logMAR acuity for E-Chart was 0.05 (SD = 0.05) and for iPad acuity was 0.04 (SD = 0.05). The improvement in mean visual acuity with the addition of the PH was 55% (0.11 vs 0.05) for E-Chart and 42% (0.07 vs 0.04) for iPad acuity while SD improved 67% (0.16 vs .05) for E-Chart and 58% (0.12 vs 0.05) for iPad acuity. The limit of agreement between logMAR iPad acuity and E-Chart was analyzed by the method of Bland and Altman,^3^ where 95% of differences will lie between plus and minus 2 SD of the mean difference (d) between the tests. Without the PH (Figure 2), for E-Chart minus iPad acuity, d = 0.04 logMAR (SD = 0.14, d-2SD = -0.25 and d+2SD = 0.32). With the PH (Figure 3), for E-Chart minus iPad acuity, d = 0.02 logMAR (SD = 0.05, d-2s = -0.08 and d+2s = 0.11).

**Figure 2.**
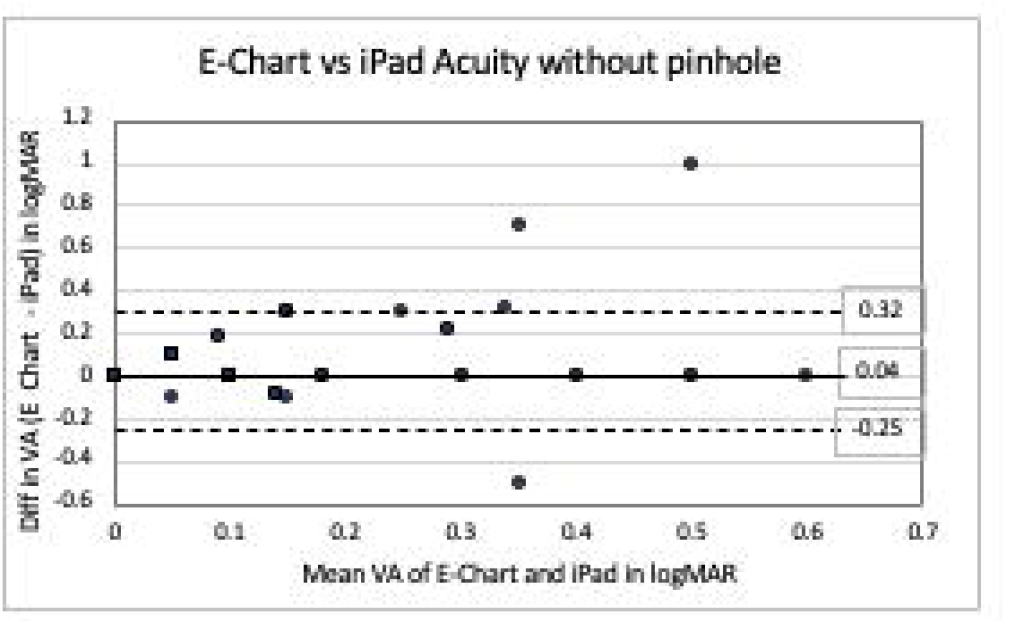
Bland-Altman plot of agreement between E-Chart and iPad visual acuities without pinhole for 63 subjects, 126 eyes.

**Figure 3.**
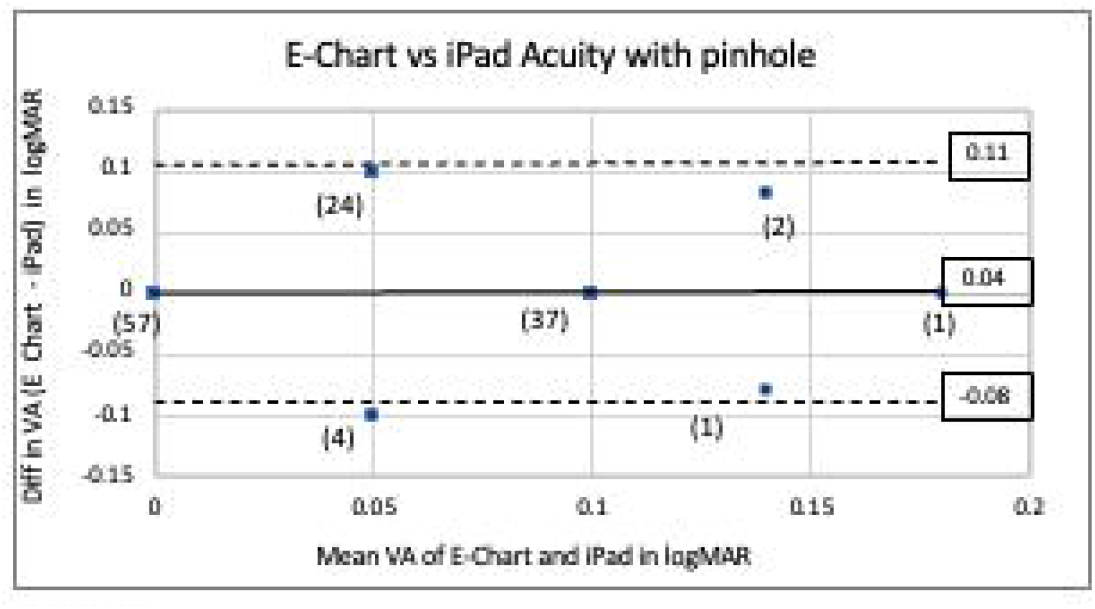
Bland-Altman plot of agreement between E-Chart and iPad visual acuities with pinhole for 63 subjects, 126 eyes, frequency (n).

Of the 204 subjects tested for brightness disparity, 2 had interocular brightness imbalance and 202 did not have brightness imbalance. When their ophthalmic status was unmasked, the first of these two subjects was known to have amblyopia and was under treatment. This child being treated for amblyopia OS had acuities measuring 20/20 OD and 20/25 OS at the time of testing. Left brightness disparity thrice measured 0.3 log unit. Colour vision was normal in each eye. The second of these two subjects was not under ophthalmic care and was referred for complete ophthalmic examination. This child was confirmed to have previously undetected left amblyopia with acuities of 20/20 OD and 20/40 OS and no visual acuity improvement with PH. Left brightness disparity twice measured 0.6 log unit and once measured 0.3 log unit. Colour vision testing suggested a blue-yellow defect OS and normal colour vision OD. A third subject who had been successfully treated for amblyopia OS had acuities measuring 20/20 in each eye. Brightness disparity testing found alternating ocular preference with endpoints of 0.3 OS, 0.3 OD, and 0.0 log units. Colour vision was normal in each eye.

Of the 204 subjects tested for colour vision, 198 students tested normal, and 6 subjects displayed a colour vision defect using the AO-HRR colour plates in an iPad. Five with a defect were bilaterally identical, classified as hereditary, with 1 female (0.83% of the females) and 4 males (4.5% of the males). The remaining subject had a monocular colour vision defect in the amblyopic eye (the child with amblyopia described above).

For brightness disparity, testing time was measured in 62 subjects. Times ranged from 12 to 63 seconds with the mean time of 32.7 seconds and standard deviation of 9.8 seconds. For iPad acuity, testing time was measured in 36 subjects. Times ranged from 41 to 188 seconds with mean of 89 seconds with standard deviation of 35.6 seconds. Some subjects began testing at the 20/60 level and others at the 20/400 level; this difference was not factored into the recording time. For colour vision testing, testing time was measured in 38 subjects. Times ranged from 17 to 95 seconds with mean of 52.8 seconds and standard deviation of 25.4 seconds.

Exit pleasure polls on a scale from 1 (boring) to 10 (fun) were taken in 60 children. The average pleasure scores were 9.7 for brightness disparity testing, 9.0 for iPad acuity, and 9.6 for colour vision testing. The younger subjects reported higher pleasure scores than the subjects between 8 and 13 years of age.

## DISCUSSION

The prevalence of amblyopia in this cohort is approximately 1%, within the reported prevalence of amblyopia worldwide between 1% and 4%^4^. Prevalence in our cohort toward the lower end of this range may reflect our recruitment process. We hypothesize that many students at this school already receive private ophthalmic care. Parents of some children with known amblyopia and other ocular conditions may have chosen to not respond to our invitation to participate in this study, possibly decreasing our measured prevalence of amblyopia.

The standard method for detecting amblyopia remains complete ophthalmic examination and measurement of best corrected acuity.^5^ In primary care and school settings, commercially available instrument-based screening devices are common.^6-10^ These devices are mostly designed to detect risk factors for amblyopia such as refractive error, strabismus, anisocoria, and media opacities rather than relative decreased acuity or amblyopia. These risk factors occur in 21%^11^ whereas amblyopia affects only 2 to 3%^1^ of the population in the United States, a disparity that may explain the inverse relationship between sensitivity and specificity of these devices according to the referral criteria chosen by the manufacturer or operator.^12^ Two more recently introduced devices, the Pediatric Vision Scanner^13^ and Diopsys^14^ objectively measure retinal birefringence and visually evoked potentials (VEP), respectively, to detect asymmetry between eyes and identify unilateral amblyopia. These devices are expensive, are not widely available in schools and pediatric offices, and are not readily applicable to telemedicine.

Why interocular brightness sense is useful to detect amblyopia is unknown. Many authors have found brightness sense useful for studying optic nerve disease. None to our knowledge state that normal brightness balance excludes disease. Inducing interocular brightness imbalance was found to severely impair hitting by major league baseball players^15^, suggesting that brightness sense influences motion stereopsis and may be evolutionarily old and conserved. A study of colour rivalry suppression in patients with ocular disease and amblyopia suggested that brightness disparity might also accompany unilateral amblyopia^16^. Our study supports this hypothesis that measurement of interocular brightness sense while playing a game on a readily available mobile graphic device, an iPad, may be a sensitive and specific method to detect unilateral amblyopia. Specialized, expensive equipment is not needed for this testing, making this methodology potentially attractive for online vision screening and for telemedicine.

Acuity testing with iPad using tumbling Es was equivalent to distance testing in our cohort and compares favorably with other methods of acuity measurement. Without a pinhole, amblyopes are not segregated from those with only refractive error. In screening children having unknown refractive errors for amblyopia, we found that adding a panoramic PH improves acuities for iPad acuity and E-Chart to a level capable of excluding amblyopia with either eye chart. Applying Bland and Altman statistics,^3^ E-Chart verses iPad acuity with PH (d= 0.02, d+2SD = 0.11, d-2SD = -0.08) showed closer agreement than when other charts^17^ were compared to E-Chart by this method: E-Chart vs HOTV (d= 0.17, d+2s = 0.37, d-2s = -0.03) and E-Chart verses Lea symbols (d= 0.15, d+2s = 0.36, d-2s = -0.07). Our limits of agreement (0.11 and -0.08) suggest that E-Chart and iPad acuity with panoramic PH can be used interchangeably. Combining measures of interocular brightness disparity and acuity with PH permits identification of unilateral amblyopia by two methods using one device.

The incidence of bilateral amblyopia has been estimated to be 0.5% and the interocular acuity difference can be very small, less than 1 lines of letters in 50% of the patients.^18^ In our study, brightness disparity testing detected the amblyope with one line of difference in visual acuity, however more studies are needed to determine the sensitivity for detecting a minimum interocular vision difference by this method. Until that sensitivity is known, both brightness disparity and acuity testing should be used for detecting amblyopia.

Video games and smartphones and tablets are ubiquitous across many societies and popular with children. This study found measures of interocular brightness disparity and visual acuity using tumbling Es useful to detect amblyopia in children as young as 3 years. The determination of interocular brightness disparity required only an average of ½ minute testing time per eye, was easy in that only 1 young subject of the 208 subjects was unable to play the “game,” and was fun, with a mean exit pleasure score of 9.7/10. The rationale that earlier screening for amblyopia leads to better outcomes is being questioned,^19^ as is the value of current vision screening in children.^20^ Outcomes were similar when treatment was immediately initiated or delayed,^21^ so screening when children are 3 years and able to play a video game remains a reasonable approach to lessening the societal burden of visual loss due to amblyopia.

Determination of brightness disparity with a graphic mobile device as demonstrated in this study is fun and easy for children and is highly sensitive and specific for detecting unilateral amblyopia. Acuity testing with spectacles or pinhole on the same device can support the results of brightness disparity measurement and help detect bilateral amblyopia. Online vision screening and telemedicine that directly measure amblyopia rather than assess risk factors may eventually displace amblyopia as number one cause of permanent vision loss in children. The screening of children in different schools and pediatric practices comparing this methodology with existing commercial devices is planned.

## Data Availability

n/a

## FIGURE LEGENDS

**FIGURES** in attached Powerpoint

## CONFLICT OF INTEREST/FUNDING

Dr Kane, Mark Gaspich, Julia Kane and Sarah Weitzman declare no conflict of interest.

Dr. Hofeldt holds a patent relating to the content of a manuscript.

This study was unfunded.

## AUTHOR CONTRIBUTIONS

All authors made the following contributions:

1. Conceived and/or designed the work that led to the submission, acquired data, and/or played an important role in interpreting the results.
2. played an important role in interpreting the results.
3. Drafted or revised the manuscript.
4. Approved the final version.
5. Agreed to be accountable for all aspects of the work in ensuring that questions related to the accuracy or integrity of any part of the work are appropriately investigated and resolved.

